# Brain Connectomics and Severity of Internalizing Symptoms in Early Adolescence Predict Severity of Suicidal Ideation in Later Adolescence

**DOI:** 10.1101/2020.11.11.20230144

**Authors:** Jaclyn S. Kirshenbaum, Rajpreet Chahal, Tiffany C. Ho, Lucy S. King, Anthony J. Gifuni, Dana Mastrovito, Saché M. Coury, Rachel L. Weisenburger, Ian H. Gotlib

## Abstract

**Background:** Suicidal ideation (SI) typically emerges during adolescence but is challenging to predict. Given the consequences of SI, it is important to identify neurobiological and psychological predictors of SI in adolescents in order to improve strategies to prevent suicide.

**Methods:** In 109 participants (61 female), we assessed psychological characteristics and obtained resting-state fMRI data in early adolescence (ages 9-13). Using graph theoretical methods, we assessed local network properties across 250 brain regions by computing measures of nodal interconnectedness: local efficiency, eigenvector centrality, nodal degree, within-module z-score, and participation coefficient. Four years later (ages 13-17), participants self-reported their SI severity. We used LASSO regression to identify a linear combination of the most important psychological, environmental, and brain-based predictors of future SI severity.

**Results:** The LASSO analysis identified a combination of 10 predictors of future SI severity (R^2^=0.23). Severity of internalizing symptoms at baseline was the strongest predictor; the remaining 9 predictors were brain-based, including nodal degree of the inferior frontal gyrus, precentral gyrus, fusiform gyrus, and inferior temporal gyrus; within-module degree of the substantia nigra and inferior parietal lobe; eigenvector centrality of the subgenual cingulate gyrus; participation coefficient of the caudal cingulate gyrus and medial amygdala.

**Conclusions:** Our findings suggest that combining network properties and earlier internalizing symptoms may improve prediction of later SI, compared to prior symptoms and other sociodemographic variables alone. Research should validate the clinical utility of these markers as predictors of suicidal thoughts.

## Introduction

Suicide is the second-leading cause of death in adolescents, resulting in approximately 5,000 adolescent deaths annually in the United States (National Center for Health Statistics (NCHS), National Vital Statistics System, National Center for Injury Prevention and Control, Centers for Disease Control and Prevention, & National Institute of Mental Health (NIMH), 2017). Further, rates of suicide among individuals ages 10-19 years have increased dramatically over the past decade in the United States (Ruch et al., 2019). Unfortunately, given their substantial heterogeneity, suicidal thoughts and behaviors (STBs) are difficult to characterize and predict (Miller & Prinstein, 2019). In individuals with psychiatric symptoms, prior STBs are strong indicators of future suicidal thoughts relative to other risk factors such as depression or anxiety, but are still weak as predictors on their own (Franklin et al., 2017). Certainly, it is important to study individuals who already have clinically relevant risk factors, such as mood disorders (Eisenlohr-Moul et al., 2018; Su et al., 2020); however, suicidal ideation (SI) is also prevalent in nonclinical and subclinical samples of community youth. Because many community youth do not seek help for their suicidal thoughts, SI can go undetected (Hawton, Saunders, & O’Connor, 2012). Because SI is a necessary precursor to suicidal behaviors (Klonsky, May, & Saffer, 2016; Turecki & Brent, 2016) it is critical that we identify factors that predict suicidal thoughts in nonclinical samples of individuals before more severe suicidal behaviors emerge.

Examining biologically-based characteristics may increase our ability to predict SI in youth who may not yet have observable symptoms. Although the literature examining neural correlates of SI in adolescents is growing, it is still much sparser than is the literature with adults (Auerbach et al., 2020; Gifuni et al., 2020). Nevertheless, several studies have now used resting-state functional magnetic imaging (rs-fMRI) to conduct either seed-based analysis or whole-brain Independent Component Analysis (ICA) to identify functional networks associated with SI in youth. For instance, greater STBs in adolescents have been associated with higher connectivity between the precuneus of the default mode network (DMN) – a network of regions consistently related to self-referential processing, including rumination (Menon, 2011) – and superior frontal gyri; however, youth with greater STBs had lower connectivity between the posterior cingulate cortex of the DMN and a cluster that included the lateral occipital cortex and fusiform gyrus (Schreiner, Klimes-Dougan, & Cullen, 2019). Similarly, depressed adolescents with history of SI had lower within-network connectivity of the ventral DMN than did depressed adolescents without SI and healthy controls (Ho et al., in press). Lower within-network connectivity of the DMN, the executive control network – regions associated with inhibitory control and decision-making (Cao et al., 2020; Menon, 2011; Uddin, 2015), and the salience network – regions that respond to emotionally salient stimuli (Seeley et al., 2007) – has been found to be associated with greater lifetime SI (Ordaz et al., 2018). Importantly, these findings suggest that disruptions in functional connections related to STBs are widely distributed across the brain.

Another approach to examining rs-fMRI connectivity patterns involves the use of graph theoretical methods, in which the brain is represented as a network (i.e., graph) composed of nodes (brain regions) and edges (connections) (Bullmore & Sporns, 2009). Using this framework, researchers are able to characterize the functional and structural organization of the whole brain on both global (network-wide) and local (nodal) levels (Rubinov & Sporns, 2010). Graph theory allows investigators to measure functional relations between nodes, even if they do not share direct anatomical projections (Honey et al., 2009). This approach allows researchers to quantify the organizational properties of specific regions in the context of the overall brain network, which may be informative for simultaneously identifying large-scale and local disruptions in network functioning that are associated with psychopathology (Bassett et al., 2008).

Although no study has used graph theory to examine the functional neural architecture associated with SI during adolescence, a few investigations have used these methods to examine STBs in adults. These studies have typically focused on differences between depressed adults with and without histories of suicide attempt (Stumps et al., 2020; Weng et al., 2019), and among ideating individuals with and without previous attempts and non-ideating individuals with depression (Kim et al., 2017). These studies have found that connections of the thalamus and superior frontal gyrus (SFG) with the rest of the brain are associated with severity of SI. Wagner et al. (2019) reported evidence of a possible association between suicidal behavior and heritable impairments in global brain functioning, characterized by weaker connections among nodes across the brain, and by reduced functional connectivity of the ventral and dorsal PFC.

While these cross-sectional studies provide insight concerning neural organization in adults who are already engaging in harmful thoughts and behaviors, it is not clear whether similar rs-fMRI connectivity patterns in young adolescents without a history of attempt or SI can *predict the severity* of suicide-related difficulties years later. Moreover, although there is emerging evidence that measures of rs-fMRI connectivity may be associated cross-sectionally with SI above and beyond clinical characteristics (e.g., age of depression onset, severity and duration of depressive symptoms, and anxious symptoms) (Ordaz et al., 2018; Schreiner et al., 2019), there is limited research examining the utility of these measures in combination with other psychological, environmental, and sociodemographic variables in predicting SI. Addressing this gap in our knowledge is especially relevant for research of young adolescents. It is possible that brain-based factors aid the prediction of SI during a developmental period in which evidence of clinical disorders has yet to be manifested at a behavioral level.

The primary goal of this study was to use graph theoretical methods to identify whether, and which, local properties of functional brain organization in combination with psychological (internalizing severity), environmental (early life stress), and sociodemographic characteristics (age, sex) in early adolescence (ages 9-13) predict the severity of self-reported SI in later adolescence (approximately 4 years later; ages 13-17). Importantly, we examined these associations in a sample recruited from the community, unselected for psychiatric disorders or suicidal history, and with no history of STBs in early adolescence. We used rs-fMRI data to compute graphic theoretical measures of the functional interconnectedness of brain regions, including efficiency, eigenvector centrality, nodal degree, within-module degree, and participation coefficient. Each of these metrics yields unique information about a region’s functional interconnectedness with the rest of the brain.

Given the nascent stage of identifying neurobiological predictors of SI, we used machine learning (ML) to examine how theoretically important psychological, environmental, and sociodemographic variables and exploratory brain-based variables (graph metrics computed across the whole brain) operate in concert to predict future severity of SI. While there are certain challenges in understanding the clinical utility of predictors identified using ML (i.e., variables that were not selected on the basis of theory but that are identified as “important” predictors of SI; Cox et al., 2020), a key advantage of ML is that it can optimize model performance across a wide range of variables regardless of their underlying distributions. Therefore, using this data-driven approach with selected variables that have a strong theoretical foundation in combination with exploratory predictors can increase precision in characterizing variability in psychopathology (Bzdok & Meyer-Lindenberg, 2018). Further, by eliminating variables from the model that to do not increase precision, the ML approach allows us to selectively identify which factors in early adolescence best predict future SI severity. Identifying the combination of neurobiological, psychological, environmental, and sociodemographic characteristics in early in adolescence that are related to the subsequent severity of SI may help guide the development of novel preventative techniques aimed at reducing the most devastating sequela of STBs: suicide attempts.

## Methods and Materials

### Participant Recruitment

We recruited 214 participants (121 biologically female) ages 9-13 years (*M=*11.38, *SD=*1.05) from the San Francisco Bay Area to participate in a longitudinal study assessing the effects of early life stress (ELS) on neurobiological development over puberty. Because participants were matched on pubertal status, boys were older than girls by an average of 7.5 months. Participants were recruited through print and online advertisements and were unselected for psychiatric disorders or suicidality history. Exclusion criteria included contraindications for MRI scan (e.g., metal implants, braces), history of major neurological disorder, intellectual delay, and non-fluent English speakers. For the current study, participants were excluded if they did not complete a functional resting-state scan at baseline or withdrew from the study (N=25) or if their functional scan data included excessive signal dropout (N=8) or movement defined by 20% of volumes>2 SD above mean framewise displacement (FD; N=8), resulting in 174 participants (101 females). At a follow-up session 3-5 years post-baseline (*M*=4.08 years), 133 participants were assessed for severity of SI, 17 of whom did not have usable scan data from baseline, 7 of whom we excluded from this analysis based on parent and child-reported history of STBs at baseline, and 1 of whom we excluded for missing data on whether they felt sleepy throughout the scan, resulting in a final sample of 108 participants (see Table 1 for participant characteristics and *Table S1* for table of medications taken at baseline). In accordance with the Declaration of Helsinki, participants and their parents provided informed written assent and consent, respectively. This study was approved by the Stanford University Institutional Review Board and all participants were compensated for their participation.

**Table 1.**
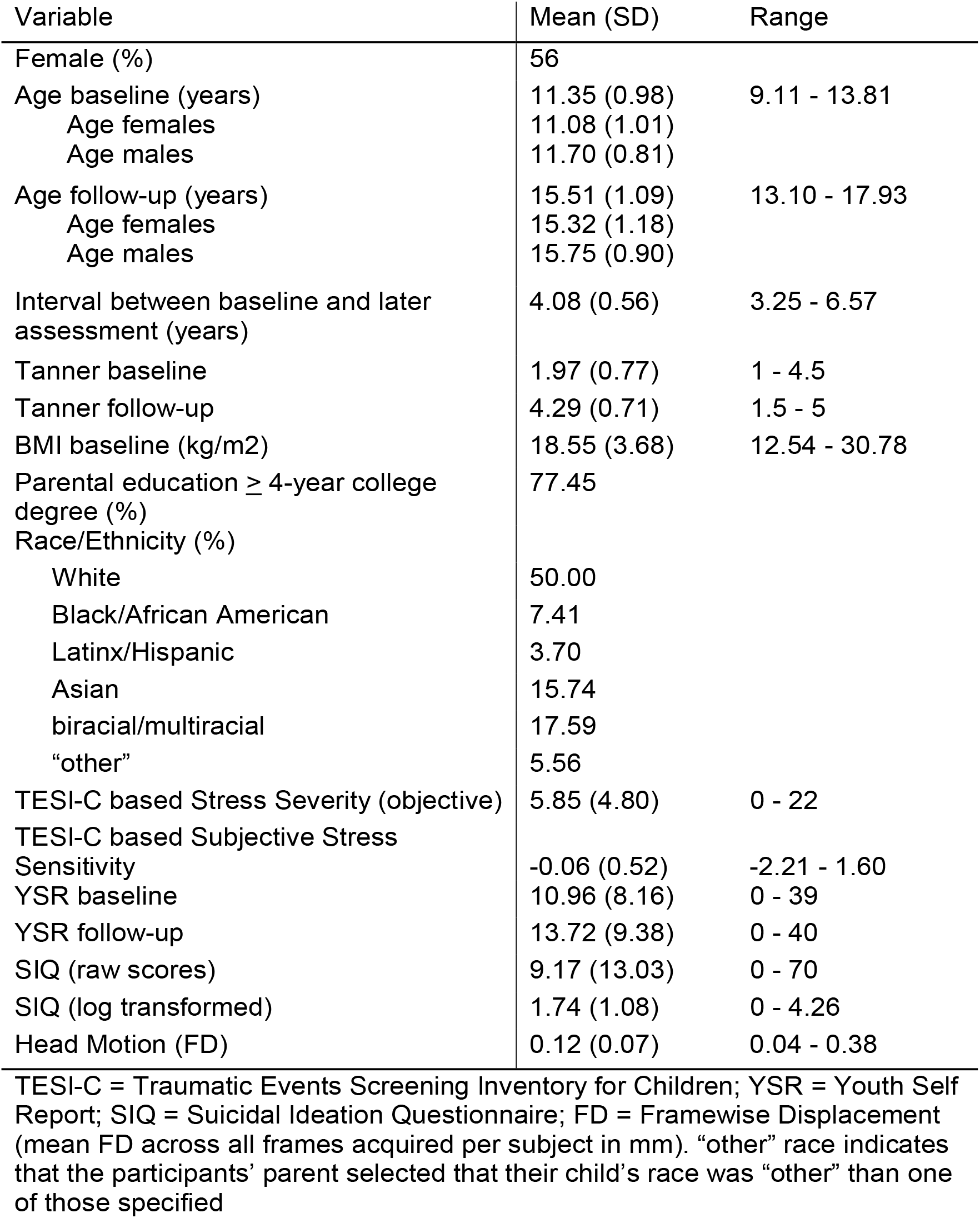
Participant Characteristics (N = 108)

### Psychological Characteristics

#### K-SADS-PL

The Kiddie-Schedule for Affective Disorders and Schizophrenia Present and Lifetime (K-SADS-PL) is a semi-structured interview used to establish presence of DSM-IV diagnoses (Kaufman et al., 1997). We asked three questions regarding STBs, involving recurrent thoughts about death, experiencing thoughts about ending their own lives, and attempting to end their own lives. These symptoms were rated as “not present,” “subthreshold,” or “threshold.” Trained interviewers administered the K-SADS-PL to children and their parents at both timepoints. For the current study we examined child- and parent-reported information of lifetime and current substance/alcohol use and STBs given that these are risk factors for future STBs (King et al., 2019). As we reported above, we excluded 7 participants who had self-reported or informant-reported threshold levels of STBs at baseline. As expected, no participant endorsed substance or alcohol use at baseline.

#### Internalizing Symptom Severity

Participants completed the Youth Self Report (YSR; Earls, Brooks-Gunn, Raudenbush, & Sampson, 2007) commonly used to assess internalizing symptoms. The YSR was administered at both time points and had high internal consistency at baseline (α=.80) and at follow-up (α=.80).

#### Suicidal Ideation

To assess dimensionality of SI, participants completed the Suicidal Ideation Questionnaire – Junior Form (SIQ-JR; Reynolds, 1988), a 15-item self-report measure of suicidal thoughts in the past month. We assessed SI severity at follow-up, the ages at which we expect SI to develop (Nock et al., 2013). Scores can range from 0-90, with higher scores indicating greater severity. The SIQ was administered at the follow-up assessment and had high internal consistency (Cronbach’s α=.97).

### Environmental and Sociodemographic Characteristics

#### Early Life Stress

To assess history of ELS, adolescents were interviewed at baseline about exposure to different types of stressful experiences using a modified version of the Traumatic Events Screening Inventory for Children (TESI-C; Ribbe, 1996). We calculated an cumulative objective stress severity score by summing the maximum objective severity scores for each type of stressor endorsed by each adolescent; this method ensured that frequent but less severe events would not be overly weighted (King et al., 2017). We also calculated a stress sensitivity score that represents participants’ cumulative subjective stress severity accounting for cumulative objective stress severity (Ho et al., 2017). See *Supplement* for information about the coding system.

#### Pubertal Status

To assess pubertal status, participants rated their developmental stage using the Tanner Staging questionnaire (Marshall & Tanner, 1969, 1970; Morris & Udry, 1980) at both baseline and follow up. This questionnaire measures developmental status based on schematic drawings of secondary sex characteristics (pubic hair and breast development for females, pubic hair and testicular development for males) on a scale from 1 (no pubertal development) to 5 (adult level of pubertal development). Within each time point, we averaged the two ratings of the secondary sex characteristics to yield a composite measure of the participants’ pubertal status. Self-reported Tanner staging has been showed to be correlated with physicians’ physical examinations of pubertal development (Coleman & Coleman, 2002; Shirtcliff, Dahl, & Pollak, 2009), and was correlated with pubertal hormones in this sample (King et al., 2020). No participant endorsed taking hormonal contraception at baseline.

#### Parent Education

As an index of socioeconomic status (SES), we assessed parental education. The parent accompanying the child indicated whether they received No GED / No High School Diploma, GED / High School Diploma, Some College, a 2-year College Degree, a 4-year College Degree, a Masters Degree, a Professional Degree (MD, JD, DDS, etc…), or a Doctorate. This was rated from 1 (No GED) – 8 (Doctorate).

#### Sleepiness

Since our instructions for the fMRI scan was to keep eyes closed, but remain awake, it is possible that sleepiness could occur. At the end of the scan, participants self-reported whether they experienced sleepiness during the scan. Most participants (N=71) experienced some feelings of sleepiness during the scan. We thus, examined sleepiness as a binary dummy-coded variable.

### fMRI Acquisition and Preprocessing

MRI scans were conducted on a GE Discovery MR750 scanner (GE Medical Systems, Milwaukee, WI) equipped with a 32-channel head coil (Nova Medical). We collected spoiled gradient echo (SPGR) T1-weighted sagittal anatomical images (repetition time [TR]=6.24ms; echo time [TE]=2.34 ms; flip angle=12°; FOV=230 mm; voxel size=0.8984 × 0.8984 × 0.9000 mm; scan time=5:15) to be used for alignment and registration of functional images and for segmenting tissue types for facilitating resting-state fMRI preprocessing. Resting-state BOLD fMRI data were acquired using aT2*-weighted echo planar imaging sequence with 37 axial slices (180 volumes, repetition time [TR]= 2.0 s; echo time [TE]=30 ms; flip angle=77°; FOV=224 mm; voxel size=3.2 mm^3^, total scan time=6:00). Participants were instructed to keep their eyes closed but remain awake. The raw functional images were quality checked prior to preprocessing by JSK and RC. The rs-fMRI data we used to compute graph metrics are derived from preprocessing performed using *fMRIPrep* 1.5.0 (Esteban et al., 2019); RRID:SCR_016216, which is based on *Nipype* 1.2.2 (Gorgolewski et al., 2011; Gorgolewski et al., 2017); RRID:SCR_002502. Preprocessing steps included skull-stripping, slice-timing and motion correction, registration to structural reference and standard pediatric template, intensity normalization, band-pass filtering, nuisance regression, and motion censoring via interpolation (see *Supplement* for complete details).

### Network Construction

#### Parcellation

To ensure sufficient spatial resolution (Craddock et al., 2012; Hallquist & Hillary, 2018), we used the Brainnetome Atlas (Fan et al., 2016) to parcellate each participant’s preprocessed structural data into 246 ROIs, plus 4 additional subregions of the basal ganglia (Keuken & Forstmann, 2015). We computed and standardized (via Fisher-z transform) the Pearson’s correlation coefficients of time-series of all pairs of regions to define the edges of the brain network, yielding a 250×250 fully connected, undirected, and weighted graph for each participant.

#### Defining edges

All negative weights in participants’ correlation matrices were set to zero to aid interpretation (Lydon-Staley et al., 2018). One challenge in conducting graph theoretical studies is deciding on a threshold to determine what constitutes a meaningful “connection” from a continuous measure of functional connectivity (Hallquist & Hillary, 2018). There is no consensus on how to threshold functional connectivity values, and most recommendations to date apply to case-control studies (Hallquist & Hillary, 2018). Consequently, we calculated graph metrics over a range of relative density thresholds (Matthews & Fair, 2015) from .10 to .20 in steps of .02. For each graph metric for each of the 250 ROIs, we summed the averages across each pair of thresholds to yield a composite threshold value (van den Heuvel et al., 2017; Hosseini, Hoeft, & Kesler, 2012; Wagner et al., 2019). See *Supplement* for more details.

#### Computation of graph metrics

All graph metrics were calculated based on each participant’s weighted correlation matrices using *GraphVar version 2*.*02* (Kruschwitz, List, Waller, Rubinov, & Walter, 2015), which uses functions from the *Brain Connectivity Toolbox* (BCT; Rubinov & Sporns, 2010). We calculated five local graph metrics per node, yielding 1250 graph-based predictors: local efficiency, eigenvector centrality, nodal degree, within-module z-score, participation coefficient because each of these metrics of functional brain organization captures different aspects of a node’s centrality and membership relative to other neighboring nodes, yielding unique information about a region’s interconnectedness with the rest of the brain (see Table 2 for definitions). We used *R version 3*.*6*.*2* (R Core Team, 2019) for all subsequent statistical analyses. See <u>JSK-github</u> for code availability.

**Table 2.** Description of Graph Metrics.

| **Metric** | **Definition** |
| --- | --- |
| Degree | Number of edges connected to that node |
| Local Efficiency | Average inverse shortest path length among the subgraph of that node; local efficiency characterizes how well information is exchanged locally when a node is removed from its neighbors (Latora & Marchiori, 2001) |
| Eigenvector Centrality | A metric of node significance in a network based on the eigenvector of the network adjacency matrix; eigenvector centrality reflects the importance of a node while giving consideration to the importance of its neighbors, assuming that more prominent nodes are linked to many other important nodes (Ballarini et al., 2018). |
| Within-module degree | Z-score of a node's within-module degree; z-scores > 2.5 denote hub status (Power et al., 2013) |
| Participation Coefficient | Measure of the distribution of a node's edges among the modules of a network; whether a node is a connector hub or a provincial hub is based on its participation coefficient. Connector hubs, which connect nodes between different modules, have higher participant coefficients; provincial hubs, which link nodes within the same module, have lower participant coefficients (Power et al., 2013). |

### Relating rs-fMRI graph-based metrics to suicidal ideation

#### SI Data Distribution and Predictors of SI

Participants scored between 0-70 on the SIQ (*M=*9.17, *SD=*13.03), with an expected low modal response of 2. Because these data were positively skewed (12.96% endorsed 0), we log-transformed the data (see *Figure S1*). We created a matrix of possible predictors of SIQ that included 1250 brain-based graph metrics obtained at baseline plus the baseline psychological (internalizing symptom severity), environmental (objective ELS severity, subjective ELS sensitivity), sociodemographic (age, sex, race, Tanner score, BMI, parent education) and other scan-related participant characteristics (the interval between baseline scan and the SIQ assessment, mean FD, and self-reported sleepiness during scan). Sex, race/ethnicity (white and person-of-color), and self-reported sleepiness during scan were dummy-coded. We imputed missing values for variables that did not have complete cases (see *Table S2*) using the Multivariate Imputation by Chained Equations (mice) package (Buuren & Groothuis-Oudshoorn, 2011) in R. This method uses the information from the other variables to predict and impute missing values. Overall, 1262 possible predictors (all continuous variables standardized) were included in the subsequent regularized regression.

#### Primary Regularized LASSO Regression

Because we had more predictors than observations and expected collinearity among many of these predictors, we conducted a regularized regression analysis to identify the combination of brain-based, psychological, environmental, and sociodemographic variables that predicted SI in later adolescence (Zou & Hastie, 2005). Specifically, we used least absolute shrinkage and selection operator (LASSO) regression, which applies the L1 penalty and thereby “shrinks” coefficient estimates of redundant variables to zero in order to identify the features that in combination yield the most predictive model (Tibshirani, 1996). After applying the appropriate penalty via cross-validation, the resulting variables will either have zero or non-zero coefficient values. The linear combination of the variables with non-zero coefficient values yields the model that best explains severity of future SI in this sample.

We used the *glmnet* package (Friedman, Hastie, & Tibshirani, 2010) in *R* to perform the regularized LASSO regression employing a full L1 penalty (i.e., α=1) and an expected gaussian distribution. We performed leave-one-out cross-validation (LOOCV) using the *cv*.*glmnet* function to determine the largest λ (i.e., hyperparameter, regularization value) associated with the smallest mean-squared error (i.e., “lambda.min”), which yields a sparse matrix of non-zero coefficients. In order to determine model performance, we computed *R*^*2*^ based on predicted and observed values. In the absence of external validation datasets, concerns of overfitting makes LOOCV a more appropriate approach than are other cross-validation techniques (e.g., validation set approaches) for identifying the optimal λ value in smaller sample sizes because in LOOCV each iteration is composed of a different training and test set, and the training set consists of almost the full dataset (James et al., 2013). In addition, LOOCV produces less bias in the test error than do other cross-validation approaches (James et al., 2013). To aid interpretation of the coefficient estimates, we computed zero-order correlations of each non-zero variable and severity of SI.

#### Supplemental Analysis of Psychological and Sociodemographic Variables

Given that sex, race, and dimensions of ELS have been found to be associated with SI (Adrian, Miller, McCauley, & Stoep, 2016; LaVome Robinson, Droege, Hipwell, Stepp, & Keenan, 2016), and because the effects of other non-brain variables in our primary LASSO may go undetected due to high penalization, we conducted a separate LASSO regression using LOOCV to determine which baseline psychological, environmental, and sociodemographic variables (sex, race, age, BMI, Tanner score, internalizing symptom severity, the interval between the time points, objective ELS severity, subjective ELS sensitivity, SES, mean FD, and reported sleepiness during scan) are associated with the severity of SI. Results and discussion of this analysis are presented in the *Supplement*, as is a full table of correlations between baseline psychological, environmental, and sociodemographic variables and severity of SI (Table S4).

#### Analysis using Internalizing Severity as an Outcome

To assess whether we captured predictors of SI specifically or internalizing symptoms more broadly, we conducted analyses with severity of internalizing symptoms as the outcome, re-running the LASSO regression replacing SIQ with YSR at follow-up, and keeping all other variables the same as in the primary analysis.

## Results

Participant characteristics are presented in Table 1. At baseline, participants were in early stages of puberty (Mean Tanner stage=1.97±0.77) and, as expected, males were on average 7.5 months older than females (*p*<0.001). In the follow-up sample four years later, when the participants were in later adolescence and more advanced puberty (Mean Tanner stage=4.29±0.71), males remained slightly older than females (*p*=0.034).

The LOOCV of the LASSO regression used to predict severity of SI yielded an optimal λ=0.18, which resulted in 10 variables with non-zero coefficients (see Table 3, Figure 1A), explaining 22.59% of the variance in severity of SI. The brain-based variables were distributed across the basal ganglia, the frontal, temporal, and parietal lobes, the cingulate gyri, and the subcortical nuclei. Higher severity of SI was predicted by lower nodal degree of the frontal gyrus – specifically the right caudal inferior frontal gyrus (β=-0.082) and the left caudal precentral gyrus (β=-0.058), followed by higher within-module degree of the right substantia nigra (β=0.058) and higher nodal degree of the temporal gyrus – specifically the left medioventral fusiform gyrus (β=0.048) and the inferior temporal gyrus (β=0.031). Additional predictors were higher eigenvector centrality of the right subgenual cingulate gyrus (β=0.028), lower within-module degree of the rostrodorsal inferior parietal lobe (β=-0.043), lower participation coefficient of the right caudal cingulate gyrus (β=-0.003), and higher participation coefficient of the right medial amygdala (β=0.0002). The only psychological self-reported predictor of SI severity was severity of internalizing symptoms at baseline (β=0.134), which was positively associated with subsequent severity of SI. None of the other environmental and sociodemographic variables had non-zero coefficients. Overall, 99.21% of the potential predictors of SI had their coefficients reduced to zero. To aid interpretation of the coefficient estimates, we computed the zero-order correlations, which revealed consistency in the direction of associations across the correlations and the LASSO results (see Table 3, Figure 1B).

**Table 3.**
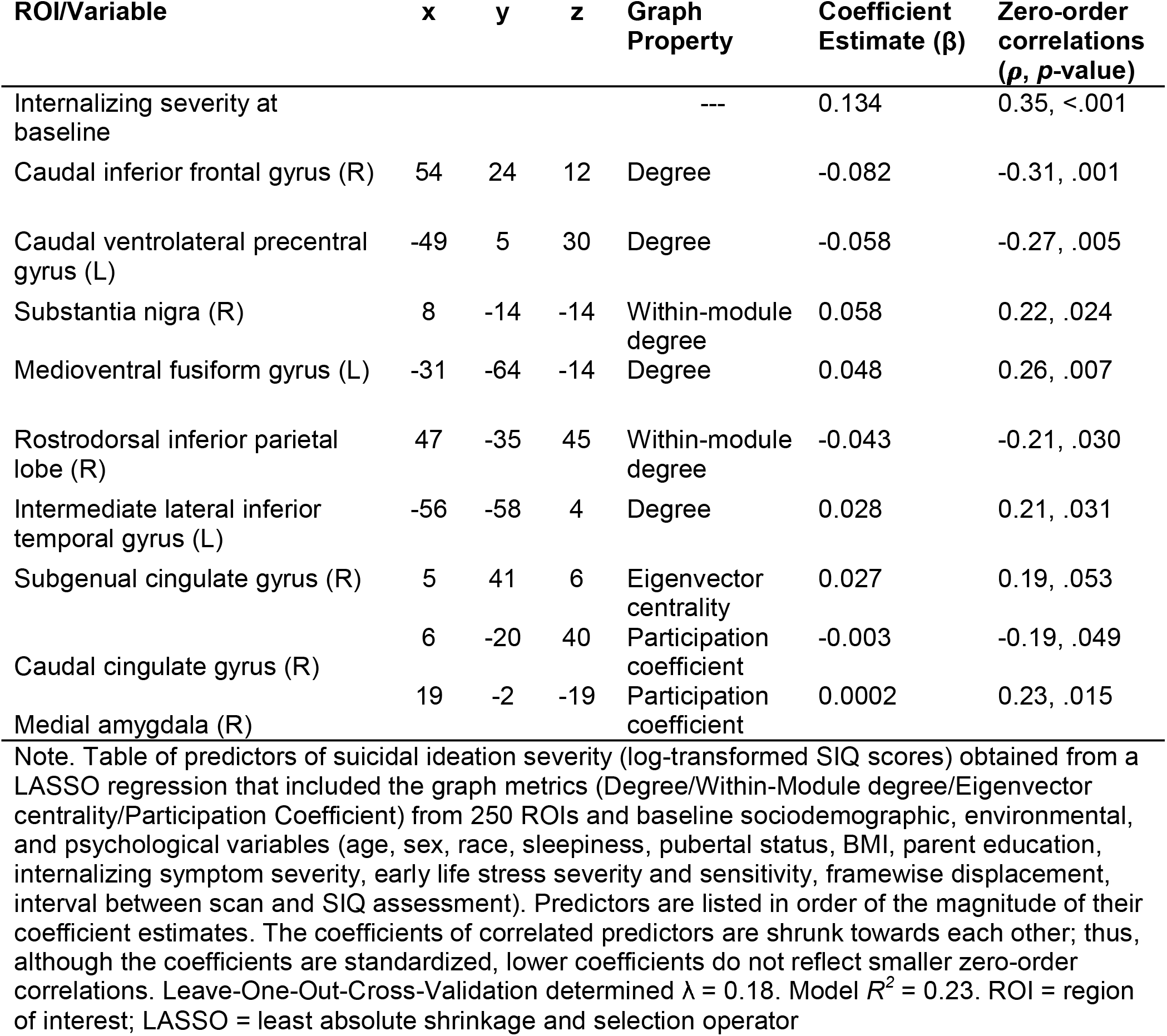
Predictors of Suicidal Ideation Severity from LASSO Regularized Regression (N=108)

**Figure 1A.**
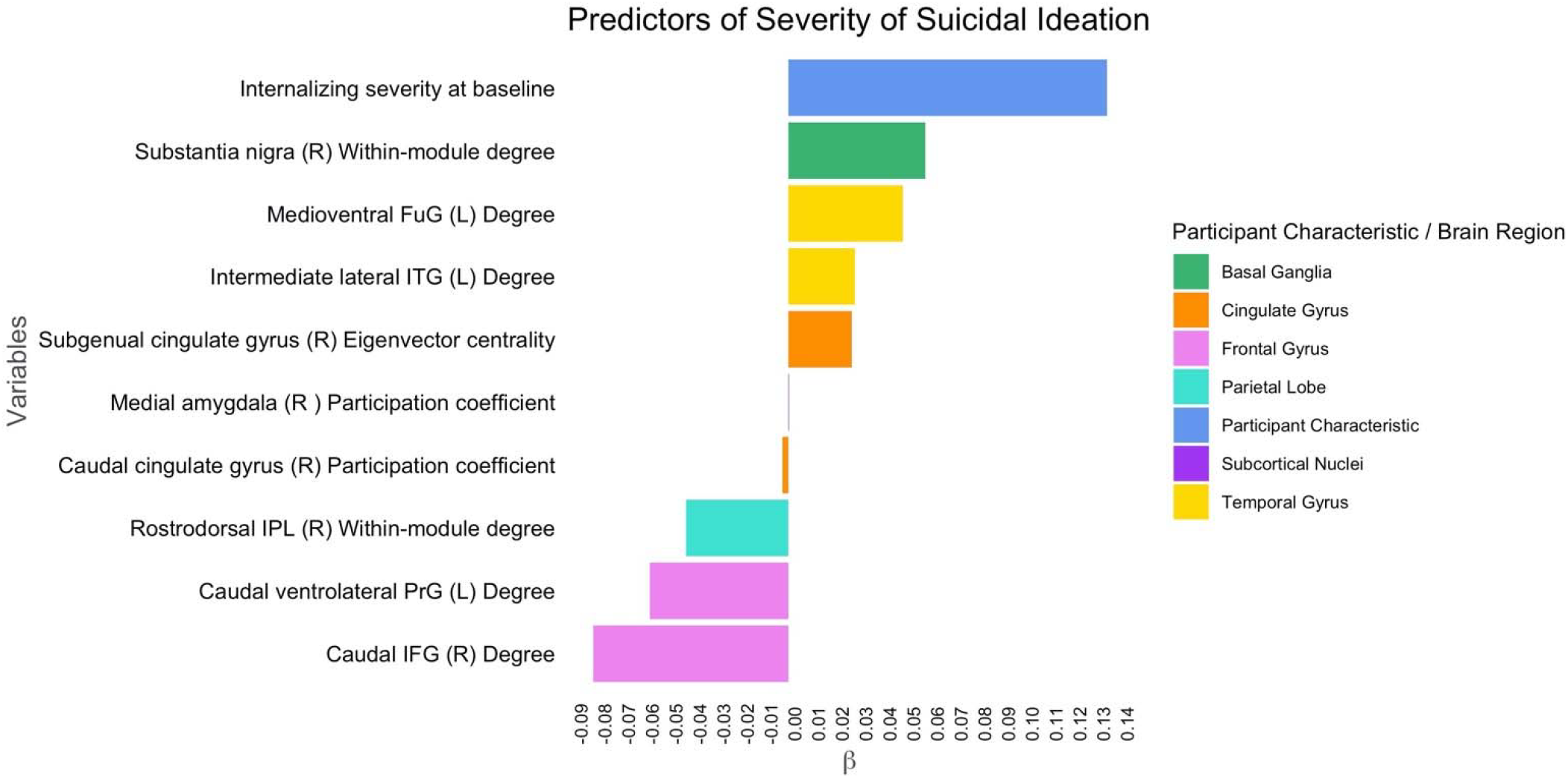
Local graph properties and psychological variables predicting severity of suicidal ideation in adolescence. Note. FuG = fusiform gyrus; ITG = inferior temporal gyrus; IPL = inferior parietal lobe; PrG = precentral gyrus; IFG = inferior frontal gyrus

**Figure 1B.**
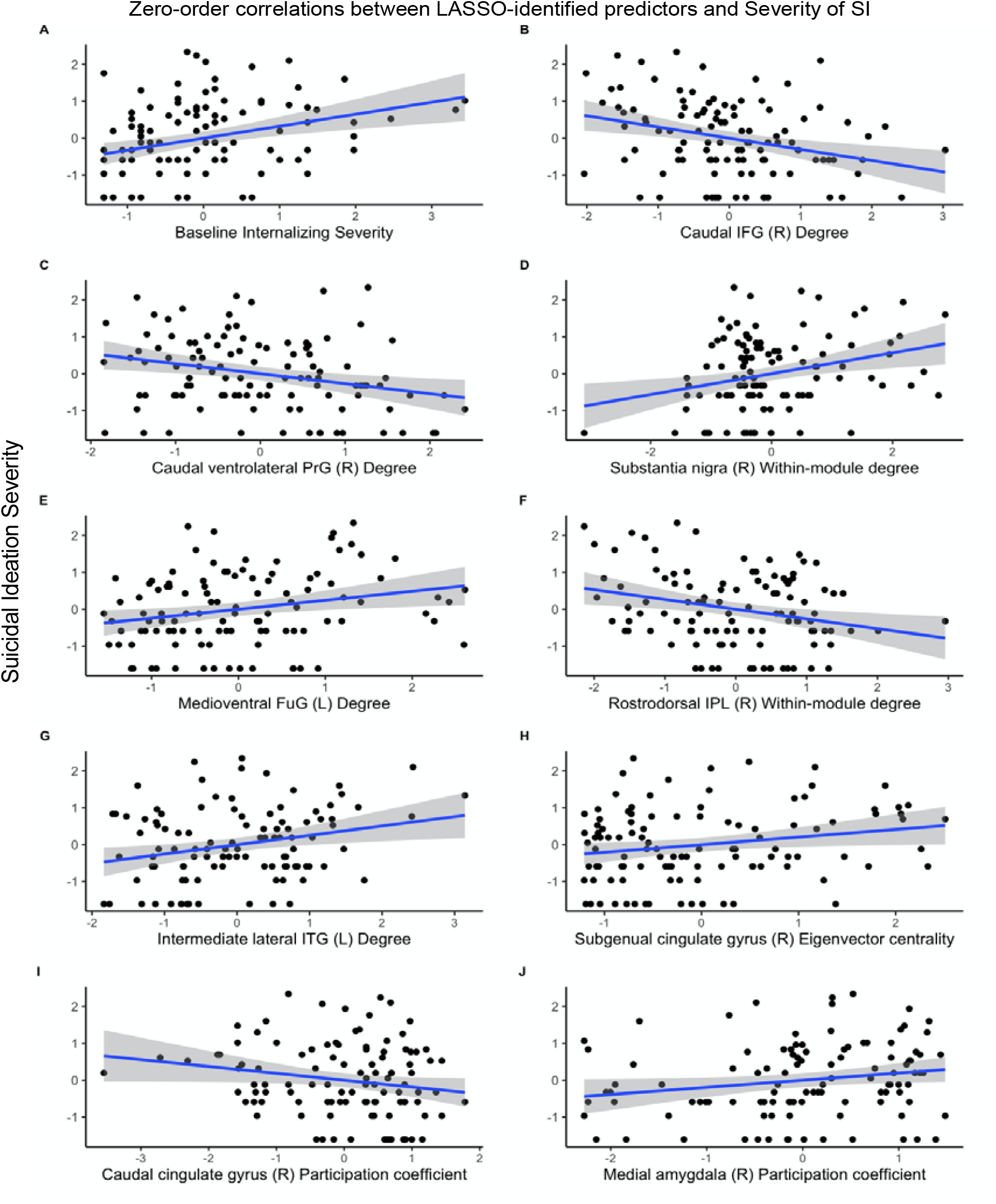
Zero-order correlations between LASSO-identified predictors and Severity of SI Note. Panels A – J represent the correlations from Table 3. SI = suicidal ideation; FuG = fusiform gyrus; ITG = inferior temporal gyrus; IPL = inferior parietal lobe; PrG = precentral gyrus; IFG = inferior frontal gyrus

In our supplementary LASSO regression of non-brain baseline variables, we observed that the combination of the severity of internalizing symptoms, sleepiness, the interval between baseline and the SIQ assessment, age at baseline, parental education, objective ELS severity, and mean FD was associated with a higher severity of SI. This model with only baseline psychological, environmental, and sociodemographic variables performed slightly worse (*R*^*2*^=0.21) compared to our primary model that included brain variables (*R*^*2*^=0.23) (results in *Table S3 and discussion* presented in *Baseline Psychological and Sociodemographic Variables, page 5 of the Supplement)*.

The LOOCV of the LASSO regression used to predict internalizing symptom severity in later adolescence yielded an optimal λ of 0.19, which resulted in a combination of 14 variables with non-zero coefficients (see Table 4 and Figure 2). Overall, we found increased interconnectedness in regions of the temporal gyrus, but reduced interconnectedness in regions of the frontal gyrus and parietal lobe in relation to internalizing severity. The participant characteristics that were also part of the model included prior internalizing severity, self-reported sleepiness during the scan, age at baseline, and sex. Overall, 98.89% of the potential predictors of internalizing symptom severity had their coefficients reduced to zero. As with our SI analysis, to help interpret the LASSO-derived coefficient estimates we computed the zero-order correlations of each predictor with internalizing symptom severity, which revealed consistency in the direction of associations across the correlations and the LASSO results (see Table 4). And as with our primary SI model, these results indicate that the severity of future internalizing symptoms is not predicted by only one brain region or one demographic characteristic; rather, the full predictive model that is the linear combination of all 14 features achieves *R*^*2*^=0.24.

**Table 4.** Predictors of Internalizing Symptom Severity from LASSO Regularized Regression (N=108)

| ROI/Variable | x | y | z | Graph Property | Coefficient Estimate ($\beta$) | Zero-order correlations ($\rho/\beta$, p-value) |
| --- | --- | --- | --- | --- | --- | --- |
| Intermediate lateral inferior temporal gyrus (L) | -56 | -58 | 4 | Eigenvector centrality | 0.132 | 0.25, .010 |
| Medioventral fusiform gyrus (R) | 31 | -62 | -14 | Degree | 0.084 | 0.22, .020 |
| **Internalizing severity at baseline** |  |  |  | --- | **0.072** | **0.35, <.001** |
| Sleepiness |  |  |  | --- | 0.069 | 0.58, .004 |
| Age at baseline |  |  |  | --- | -0.051 | -0.20, .036 |
| Orbital area 12/47 (R) | 40 | 39 | -14 | Participation coefficient | -0.037 | -0.24, .011 |
| Sex |  |  |  | --- | 0.031 | 0.54, .005 |
| **Caudal inferior frontal gyrus (R)** | **54** | **24** | **12** | **Degree** | **-0.027** | **-0.27, .004** |
| Caudal parahippocampal gyrus (L) | -25 | -25 | -26 | Local efficiency | 0.018 | 0.19, .048 |
| **Rostrodorsal inferior parietal lobe (R)** | **47** | **-35** | **45** | **Within-module degree** | **-0.017** | **-0.19, .055** |
| Entorhinal cortex (R) | 19 | -10 | -30 | Eigenvector centrality | 0.016 | 0.23, .016 |
| Ventral inferior frontal gyrus (L) | -52 | 13 | 6 | Eigenvector centrality | -0.011 | -0.21, .032 |
| Caudal inferior frontal gyrus (R) | 54 | 24 | 12 | Within-module degree | -0.007 | -0.25, .009 |
| Posterior parahippocampal gyrus (R) | 30 | -30 | -18 | Local efficiency | 0.005 | 0.15, .130 |

Note. Table of predictors of internalizing symptom severity obtained from a LASSO regression that included the graph metrics (Degree/Within-Module degree/Eigenvector centrality/Participation Coefficient) from 250 ROIs and baseline sociodemographic, environmental, and psychological variables (age, sex, race, sleepiness, pubertal status, BMI, parent education, internalizing symptom severity, early life stress severity and sensitivity, framewise displacement, interval between scan and SIQ assessment). Leave-One-Out-Cross-Validation determined $\lambda = 0.19$ . Model $R^2 = 0.24$ . Predictors are listed in order of the magnitude of their coefficient estimates. The coefficients of correlated predictors are shrunk towards each other; thus, although the coefficients are standardized, lower coefficients do not reflect smaller zero-order correlations. **Bolded variables indicate overlap with predictors of suicidal ideation.** ROI = region of interest; LASSO = least absolute shrinkage and selection operator

**Figure 2.**
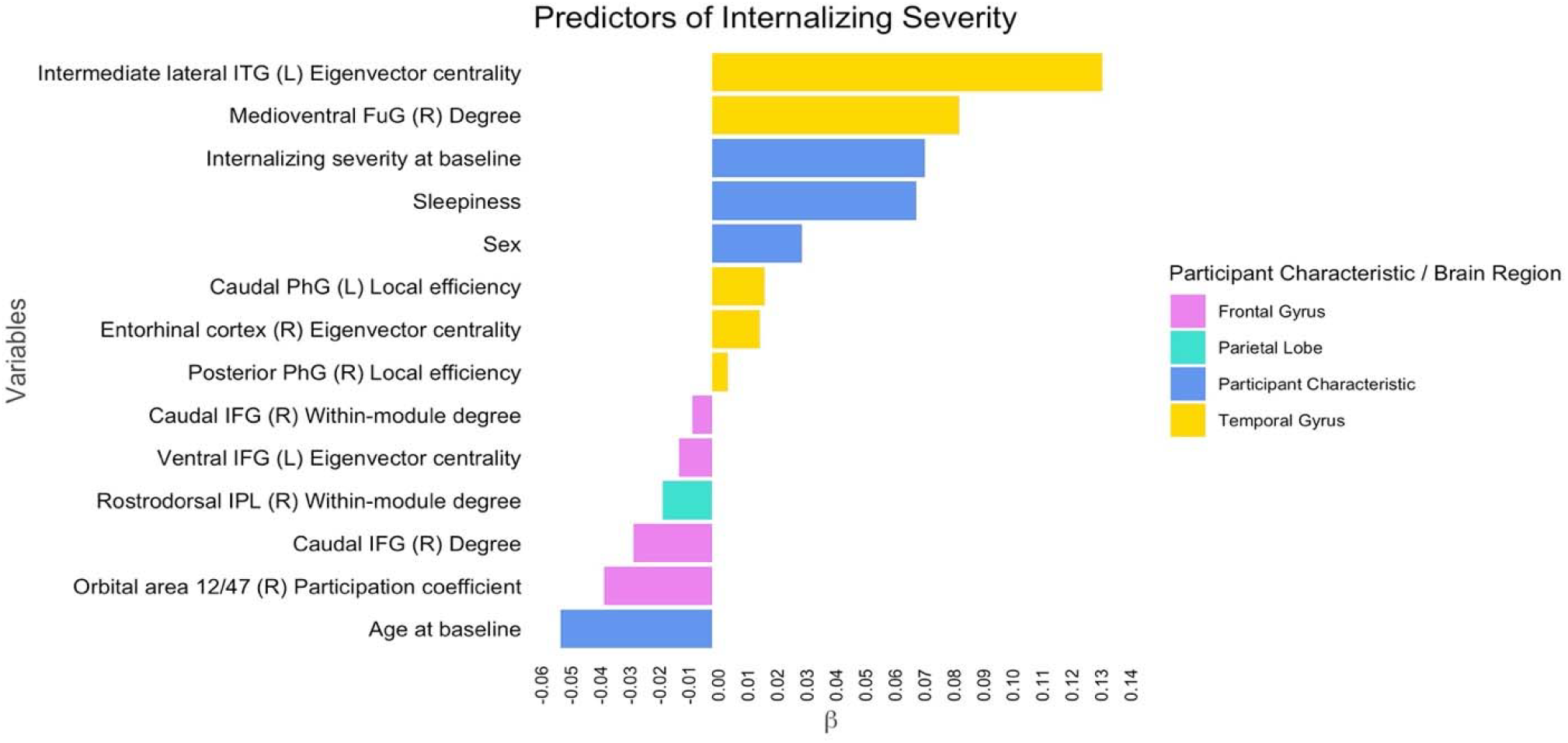
Local graph properties and psychological variables predicting severity of internalizing symptoms in adolescence. ITG = inferior temporal gyrus; FuG = fusiform gyrus; PhG = parahippocampal gyrus; IFG = inferior frontal gyrus; IPL = inferior parietal lobe

## Discussion

Recent meta-analyses have reported relatively weaker effect sizes of clinical and sociodemographic characteristics than of prior STBs as predictors of future SI (Franklin et al., 2017); however, STBs can go undetected in samples of community youth (Hawton et al., 2012). Here, we examined whether interconnectedness of brain regions, measured with rs-fMRI in an unselected sample of early adolescents predicts the severity of SI in later adolescence. We also included in the analyses psychological, environmental, and sociodemographic characteristics measured in early adolescence to examine which combination of variables that best predict future severity of SI. A machine learning approach allowed us to elucidate which theoretically important psychological (internalizing severity) and environmental (early life stress) variables, as well as more exploratory brain-based variables (graph metrics computed across the whole brain) operate in concert to predict future severity of SI. We found that several brain-based graph metrics predicted subsequent severity of SI, including some regions that have been identified in previous studies as being associated with STBs, such as the precentral gyrus (Sublette et al., 2013), inferior frontal, temporal, cingulate gyri (Auerbach et al., 2020; Schmaal et al., 2020), and subcortical regions (amygdala) (Alarcón et al., 2019), as well as other brain structures that have not previously been linked with SI (substantia nigra).

In temporal and mesial temporal regions, more connections of the left lateral inferior temporal gyrus, the left fusiform gyrus, and higher participation of the right medial amygdala with the rest of the brain were related to greater future severity of SI. Few fMRI studies have found associations between SI and the lateral inferior temporal gyri, but some structural MRI studies have found reduced volume of this structure in adults diagnosed with borderline personality disorder who have attempted suicide (Soloff et al., 2012). In task-based fMRI studies those with a history of attempt show reduced activation in the left fusiform gyrus when viewing neutral compared to angry emotional faces (Pan et al., 2013) and reduced regional homogeneity (lower synchronization of resting-state activity; Yuan et al., 2013) in this region. Depressed adolescents with high SI and history of attempts had stronger functional connectivity between the amygdala and other prefrontal and cingulate regions compared to those with lower SI and healthy controls (Alarcón et al., 2019). Within the frontal gyrus, we found reduced interconnectedness, particularly lower degree in left precentral gyrus in relation to future severity of SI. Studies using PET imaging have reported that the relative metabolic rate of glucose was lower in the precentral gyri in adults who attempted suicide than in those without suicide attempts (Sublette et al., 2013). Importantly, these previous studies have examined those with a history of attempt; thus, it is important that future studies examine explicitly and systematically whether the connectedness of these brain regions differs depending on the trajectory of adolescents’ suicide-related thoughts and behaviors.

The graph metrics we identified are important in elucidating the organizational properties of functional connectivity previously implicated in SI. In summary, we found here that a lower number of connections (degree) of areas in the frontal gyrus and a higher number of connections of areas in the temporal gyrus predict future severity of SI. In addition, a higher number of connections of the substantia nigra with its surrounding regions (within-module degree) was associated with increased severity of SI. The majority of dopaminergic neurons in the brain originate in the substantia nigra (González-Hernández & Rodríguez, 2000; Zhou & Lee, 2011), which is typically studied in the context of motor (Utter & Basso, 2008) and reward-related (Richter et al., 2020) behavior, but is also targeted in deep brain stimulation. Indeed, one case study found that acute stimulation of this region evoked an acute depressive episode with thoughts of “not wanting to live” (Blomstedt et al., 2008). Consistent with our findings, the degree to which this region shares connections within the basal ganglia may also participate in the biological characterization of SI.

We also identified a higher eigenvector centrality of the right subgenual cingulate cortex (sgCG) associated with SI, which indicates that the connectivity of this region with other highly connected regions is associated with higher severity of SI. Previous findings of associations between the sgCG and SI have been mixed. For instance, in individuals with treatment-resistant depression, there was no evidence of an association between regional glucose metabolism of the sgCG and SI (Ballard et al., 2015); however, other studies have found that inflammation in this region is higher in individuals with SI than in those without SI (Holmes et al., 2018). In a recent study examining graph metrics in depressed adolescents, organizational properties other than eigenvector centrality of the right sgCG were associated with depression severity (Gabbay et al., 2020); in our sample, however, we did not find this region to be included in the model predicting increased internalizing severity. Within the cingulate cortex, we also found reduced participation of the right caudal cingulate gyrus, indicating that this region participates less in networks of other regions compared to its own network of regions. The caudal cingulate gyrus, sometimes referred to as the dorsal or midcingulate region of the cingulate (Stevens et al., 2011) and has been implicated in studies where patients have completed suicide, where higher regional cerebral blood flow was associated with those who later completed, compared to regional cerebral blood flow in patient controls with depression (Amen et al., 2009). The cingulate cortex, particularly the sgCG has been implicated across several mood disorders (Drevets, Savitz, & Trimble, 2008); however, our findings that elucidate the connectedness of the cingulate cortex that may aid future work examining specificity of this region in relation to the processes that lead to a greater severity of SI.

We found both common and distinct brain-based predictors of SI compared to internalizing symptom severity. The associations between interconnectedness of brain regions and internalizing symptom severity were generally located in frontal, parietal and temporal lobes and, thus, less widespread than the associations between interconnectedness and severity of SI, which were distributed across frontal, parietal, and temporal lobes, as well as the basal ganglia, cingulate gyrus, and subcortical regions. Importantly, the gyri that were implicated in predicting severity of both internalizing symptoms and SI had the same direction of association – that is, interconnectedness of frontal gyrus regions were negatively correlated with future severity of internalizing problems and of SI, whereas interconnectedness of temporal gyrus regions were positively correlated with these features.

The specific frontal and temporal gyri regions implicated in SI were distinct from internalizing, except for reduced nodal degree of the right inferior frontal gyrus and reduced within-module degree of the right rostrodorsal inferior parietal lobe, which were shared. In their recent review, Schmaal et al. (2020) suggest that the frontal gyrus, specifically the inferior region, facilitates the transition from suicidal thoughts to behaviors because it is involved in cognitive flexibility and behavioral decision-making (Schmaal et al., 2020), and increased functional connectivity of the right inferior frontal gyrus has been associated with depression (Connolly et al., 2013; Rolls et al., 2020). In addition, the right inferior frontal gyrus has been consistently associated with inhibitory control, such that reduced activation is associated with reduced inhibition (Rubia et al., 2005). Within the parietal lobe, there was a lower number of connections of the right inferior parietal lobe with its surrounding regions may be contributing to both higher severity of SI and internalizing symptoms. This organizational property of the right inferior parietal lobe is consistent with research showing that individuals with depression, SI, and suicidal plans have reduced regional glucose metabolism in the right inferior parietal lobe compared to those with SI only and to healthy controls (van Heeringen et al., 2017). More research is needed to understand whether reduced regional glucose metabolism is related to a lower number of connections to its surrounding regions. Based on these prior studies and our results, it could be that reduced activation and reduced interconnectedness are characteristics of behaviors that are shared across depressive and SI symptomologies.

Importantly, we did not identify some regions that have been implicated in previous studies (see Schmaal et al., 2020, and Auerbach et al., 2020, for reviews). For example, we did not find evidence that the thalamus predicted the future severity of SI. Previous studies have shown that depressed individuals with a suicide attempt history have increased connectivity of the thalamus relative to depressed individuals without a history of attempt (Jung et al., 2020); in contrast, however, Pan et al. (2011) found decreased activity of the right thalamus scanned during high-risk decisions in depressed adolescents who had attempted compared to depressed adolescents who had not attempted suicide. It is possible that this region is more strongly related to suicidal behaviors experienced in depression than to suicidal thoughts.

There was only a slight decrease (1.19%) in model fit by excluding brain-based variables from our model with psychological, environmental, and sociodemographic characteristics. Although, the change in model fit was modest, our analyses show that incorporating neurobiological factors has some added value in predicting the severity of SI in adolescents— particularly before individuals engage in their first suicide gesture. Identifying neurobiological predictors of SI in combination with psychological, environmental, and sociodemographic variables may help guide research into the cognitive and affective processes that underlie the emergence of SI, which may then inform the development of prevention and early-intervention based programs.

Contrary to our hypothesis and to findings in other studies of a relation between adversity and STBs in samples of adolescents (Khan et al., 2015) we did not find that ELS severity was related to later severity of SI in our primary model that incorporated the brain. We took a continuous approach to measuring ELS in this study, assessing a wide range of experiences, including both relatively normative stressors (e.g., moving, parental divorce) and highly adverse events (e.g., physical abuse, sexual assault); approximately 12% of our sample was exposed to these severe events (King et al., 2019). It is possible that ELS severity was not identified in our model because our main outcome was severity of SI and not attempt (Stewart et al., 2019). We may have also found stronger effects of ELS had more children in our sample been exposed to more highly severe adverse events (see *Supplemental Details* for more discussion on psychological, environmental and sociodemographic variables).

We should note four limitations of this study. First, we did not obtain information about family history of STBs. Given previous findings of a significant genetic component predicting suicide attempts (Ruderfer et al., 2019), obtaining family history of STBs may have yielded important information about subsequent severity of SI. Additionally, recent evidence from the Adolescent Brain Cognitive Development Study reported that high family conflict low parental monitoring was associated with SI (DeVille et al., 2020). Incorporating this information into future studies will be important. Second, the distribution of SI at follow-up was skewed, with most participants endorsing low severity of SI. A distribution with more severe ideation may have yielded different results; future studies will benefit from including a wider range of SI. Third, we used LOOCV to test the reliability of our results; internal CV can help reduce overfitting to some extent, but overfitting of the tuning parameter used to train the model can still occur (Hosseini et al., 2020). Our findings should be replicated using independent samples for CV (Hosseini et al., 2020). Finally, our resting-state scan was 6 minutes long. Although longer scan times can improve the reliability of functional connectivity estimates (Birn et al., 2013; Termenon et al., 2016), estimates have been found to be reliable in scans as short as 5 minutes (Andellini et al., 2015; Van Dijk et al., 2010).

Despite these limitations, the present study underscores the importance of examining distributed functional architecture of the brain in early adolescence to predict SI severity in later adolescence, a time when suicidal thoughts dramatically increase (Nock et al., 2013). A strength of our investigation is that we recruited a sample of adolescents with minimal psychiatric histories and were able to identify several neurobiological and psychological factors that predicted the severity of SI 4 years later. Given that we identified organizational properties of regions that are distributed across the brain, it is important that future work replicate our data-driven approach, interrogating regions of the brain that might be missed otherwise. Combined with previous studies, our findings may help guide future neuroimaging research focused on understanding the neurobiological associations with the cognitive and affective processes that make individuals more prone to suicidal thoughts before the emergence of more severe suicidal behaviors. Finally, although we focused in this study on temporally distal risk factors for suicidal thoughts, gaining a better understanding of how the brain is associated with the transition from suicidal thoughts to behaviors is an important direction for future research.

## Data Availability

Data will be made available upon request.

https://github.com/jackie-schwartz/brain_connect_and_si

## Acknowledgments

We thank Michelle Sanabria, Johanna Walker, Holly Pham, and M. Catalina Camacho for their assistance with data collection and organization. Finally, we thank the participants and their families for participating in this study.

## Author Contributions

J.S.K., T.C.H., L.S.K., and I.H.G. designed research; D.M., S.M.C., and R.L.W. helped perform research; J.S.K., R.C., and T.H.C. planned analyses; J.S.K. analyzed data; J.S.K., R.C., T.C.H., and L.S.K. discussed results; J.S.K., R.C., T.C.H., L.S.K. A.J.G., and I.H.G. wrote paper.

## Supplemental Information

### Method

#### Early Life Stress Interview

For each type of stress that the participant endorsed, interviewers followed up with specific questions to characterize the severity of the stressful experience. As described in King et al. (2017, 2019), a panel of three coders, blind to the children’s subjective severity ratings and reactions and behaviors during the interview, then rated the objective severity of each type of stressor endorsed using a modified version of the UCLA Life Stress Interview coding system (Rudolph et al., 2000). Coders made objective severity ratings on a 5-point scale with an Intraclass Correlation Coefficient of 0.99 (King et al., 2017).

#### fMRI Preprocessing

Preprocessing was implemented using *fMRIPrep* (Esteban et al., 2019).

##### Anatomical data preprocessing

The T1-weighted (T1w) image was corrected for intensity non-uniformity with N4BiasFieldCorrection (Tustison et al., 2010), distributed with ANTs 2.2.0 (RRID:SCR_004757; Avants, Epstein, Grossman, & Gee, 2008), and used as an anatomical reference throughout the workflow.

The T1w-reference image was then skull-stripped with a *Nipype* (Gorgolewski et al., 2017) implementation of the antsBrainExtraction.sh workflow (from ANTs), using OASIS30ANTs as the target template. Brain tissue segmentation of cerebrospinal fluid (CSF), white-matter (WM) and gray-matter (GM) was performed on the brain-extracted T1w using *FAST* (from FSL 5.0.9, RRID:SCR_002823) (Zhang, Brady, & Smith, 2001). Brain surfaces were reconstructed using *recon-all* (FreeSurfer 6.0.1, RRID:SCR_001847) (Dale, Fischl, & Sereno, 1999), and the brain mask estimated previously was refined with a custom variation of the method to reconcile ANTs-derived and FreeSurfer-derived segmentations of the cortical gray-matter of Mindboggle (RRID:SCR_002438) (Klein et al., 2017). Volume-based spatial normalization to standard space (*MNI’s unbiased standard MRI template for pediatric data from the 4*.*5 to 18*.*5y age range* [RRID:SCR_008796; TemplateFlow ID: MNIPediatricAsym) was performed through nonlinear registration with antsRegistration (ANTs 2.2.0), using brain-extracted versions of both T1w reference and the T1w template.

##### Functional data preprocessing

First, a reference volume and its skull-stripped version were generated using a custom methodology of *fMRIPrep*. Based on the estimated susceptibility distortion, an unwarped BOLD reference was calculated for a more accurate co-registration with the T1w reference. Head-motion parameters (transformation matrices, and six corresponding rotation and translation parameters) with respect to the BOLD references were estimated using *MCFLIRT* (from FSL 5.0.9) (Jenkinson, Bannister, Brady, & Smith, 2002) before applying any spatiotemporal filtering. BOLD runs were slice-time corrected using *3dTshift* from AFNI 20160207 (RRID:SCR_005927) (Cox & Hyde, 1997). Following slice-time and motion correction, the BOLD time-series are resampled onto their original, native space using the head-motion correction and susceptibility distortion correction mappings from the previous steps, which is done using a one-shot interpolation process. After slice-time and motion correction, the BOLD reference was used to co-register to the T1w reference using *bbregister* (FreeSurfer) which implements boundary-based registration (Greve & Fischl, 2009) using rigid-body transformation. The co-registered BOLD time-series were then resampled into standard space (MNIPediatricAsym) and to surfaces on the *fsaverage5* atlas space.

Several confounding time-series regressors were calculated based on the *preprocessed BOLD*: framewise displacement (FD), the change in signal intensity across each volume (DVARS) (Power et al., 2014) and three region-wise global signals. FD and DVARS are calculated for each functional run, both using their implementations in *Nipype* (following the definitions by Power et al. 2014) (Power et al., 2014). In addition to three global signals extracted within the CSF, the WM, and the whole-brain masks, we also included a set of physiological regressors to allow for component-based noise correction, *CompCor* (Behzadi, Restom, Liau, & Liu, 2007). Principal components are estimated after high-pass filtering the *preprocessed BOLD* time-series (using a discrete cosine filter with 128s cut-off) for the two *CompCor* variants: temporal (tCompCor) and anatomical (aCompCor). tCompCor components are then calculated from the top 5% variable voxels within a mask covering the subcortical regions. This subcortical mask is obtained by heavily eroding the brain mask, which ensures it does not include cortical GM regions. For aCompCor, components are calculated within the intersection of the aforementioned mask and the union of CSF and WM masks calculated in T1w space, after their projection to the native space of each functional run (using the inverse BOLD-to-T1w transformation). Components are also calculated separately within the WM and CSF masks. For each CompCor decomposition, the *k* components with the largest singular values are retained, such that the retained components’ time series are sufficient to explain 50% of variance across the nuisance mask (CSF, WM, combined, or temporal). The remaining components are dropped from consideration. The 6-parameter head-motion estimates calculated in the correction step were also placed within the corresponding confounds file. The confound time-series derived from head motion estimates and global signals were expanded with the inclusion of temporal derivatives and quadratic terms for each (Satterthwaite et al., 2013). Frames that exceeded a threshold of 0.5 mm FD or 1.5 standardized DVARS were annotated as motion outliers. All resamplings can be performed with *a single interpolation step* by concatenating all the pertinent transformations (i.e. head-motion transform matrices, susceptibility distortion correction when available, and co-registrations to anatomical and output spaces). Gridded (volumetric) resamplings were performed using antsApplyTransforms (ANTs), configured with Lanczos interpolation to minimize the smoothing effects of other kernels (Lanczos, 1964). Non-gridded (surface) resamplings were performed using mri_vol2surf (FreeSurfer). For a full description of the preprocessing pipeline, including the workflow utilized for each preprocessing step, please refer to https://fmriprep.org/en/stable/workflows.html.

##### Post-fMRIPrep

Following preprocessing in fMRIPrep, the first 6 frames were discarded to allow the MR signal to achieve T1 equilibrium. We then conducted nuisance regression (nuisance regressors included framewise displacement, 6 motion parameters of translation and rotation, and their first and second derivatives, and aCompCor components 0-5) followed by temporal band-pass filtering (.01 - .1 Hz) to the resting-state data. Post-processing steps included intensity normalization of the time series (sum of squares=1), and motion-censoring (censored values are replaced by interpolated neighboring in time non-censored values) for frames that exceed 0.5mm FD.

#### Defining Edges

For each graph metric for each of the 250 ROIs, we computed the area under the curve with respect to ground (AUC_g_) to summarize the relative density thresholds. We multiplied the difference between each threshold (.02) by the average between each threshold pair to yield a composite for each the threshold pairs and then summed these composites to yield the total AUC_g_, which is then used for subsequent analyses. See scripts 1 – 5 <u>here</u> for code.

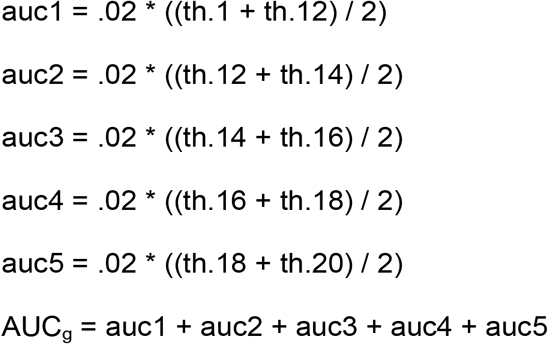

### Supplemental LASSO Analysis of Baseline Psychological, Environmental and Sociodemographic Variables

We conducted a separate LASSO regression using LOOCV to determine which baseline psychological and sociodemographic variables (sex, race, age, BMI, Tanner score, internalizing symptom severity, the interval between the time points, objective ELS severity, subjective ELS sensitivity, socioeconomic status, mean FD, and reported sleepiness during scan) are associated with the severity of SI. Consistent with our primary LASSO regression, we found that severity internalizing symptoms was the strongest predictor. Additional predictors included sleepiness during the scan, the years between the baseline assessment and SIQ assessment, age at baseline, parent education, severity of objective ELS, and mean FD (see Table S3).

We expected sex to be part of the linear combination of variables predicting severity of SI. Some research has reported that sex differences are more common in self-harming behaviors than in SI (Hawton, Saunders, & O’Connor, 2012; Mars et al., 2019; Oquendo et al., 2007). In contrast, however, there are also robust findings of a higher prevalence of ideation and attempts in girls than in boys (Fox, Millner, Mukerji, & Nock, 2018). Sex did not appear as a predictor in our primary LASSO analysis, possibly due to the penalization level. There is consistent evidence of sex differences in levels of internalizing symptoms during adolescence, particularly after puberty (Thapar, Collishaw, Pine, & Thapar, 2012), which could be why we observed this variable as part of the predictive model for future internalizing symptom severity. We did not find that pubertal stage, or BMI, at baseline predicted severity of SI. However, there are several dimensions of puberty, for instance pubertal timing (i.e., the age of pubertal onset relative to other same-aged same-sex peers) that has been found to predict severity of SI (Patton & Viner, 2007). We cannot make conclusions about pubertal timing in our sample because we recruited girls and boys matched on pubertal status and not on age, which led us to excluding girls who started puberty early relative to their peers. We did find that adolescent age and longer intervals between the baseline assessment and the SIQ assessment were positive predictors of future severity of SI in combination with our other non-brain variables. Previous research has reported that SI peaks at age 13 in girls and then decreases, whereas for boys the peak may be earlier (Adrian, Miller, McCauley, & Stoep, 2016). Future research with sufficient power to examine sex differences should consider examining pubertal- and age-related trajectories of SI. In our model excluding brain features, we found that severity of ELS was associated with the severity of future SI.

It is well recognized that exposure to stressful life events is a potent risk factor for the development of suicidal thoughts and behaviors in adolescents (Dykxhoorn, Hatcher, Roy-Gagnon, & Colman, 2017; Miller & Prinstein, 2019). It is possible that our sample on average did not experience high enough levels of stress to detect an effect in our primary analysis that takes the brain into consideration.

We also recruited a convenience sample, which represented the local demographic composition of the Bay Area and thus was not designed to examine race/ethnicity in relation to SI. Although our study did not detect race/ethnicity as a predictor of SI, previous literature has reported differences in SI and attempts across different races and ethnicities (Ivey-Stephenson, 2020; LaVome Robinson, Droege, Hipwell, Stepp, & Keenan, 2016). It is important for future research to interrogate trends of SI and how thoughts may transition to behaviors across races/ethnicities during adolescence. We did find that higher parent education was part of our non-brain model predicting severity of SI, which is contrary to findings from previous research (Hallfors et al., 2004). It could be that there is an association at lower and higher levels of parental education; however, given our sample was mostly graduated from college, it may be more difficult to assess whether lower levels of parental education would be associated with severity of SI.

Interestingly, we found that self-reported sleepiness during the scan was associated with future severity of SI. Poor sleep quality has been associated with SI (Littlewood et al., 2019); however, it is unclear if those who felt sleepy during the scan also had sleep problems in general. It is important for researchers to consider collecting data on whether participants felt sleepy during fMRI studies when examining psychological problems that are comorbid with sleep problems. Mean FD was also a predictor in the LASSO model with non-brain variables, where lower levels of average motion were associated with higher severity of SI. It is possible that those who were sleepier were also less likely to move throughout the scan.

**Table S1.** Medications at Baseline.

| **Type** | **Generic (Brand)** | **N** |
| --- | --- | --- |
| Antihistamine | Lortadine (Claritin), Cetirizine (Zyrtec), Fexofenadine (Allegra), un-named allergy medication | 6 |
| Anti-inflammatory | Montelukast (Singulair) | 1 |
| Steroid or Corticosteroid | Fluticasone Propionate (Flonase), formoterole and mometasone (Dulera) | 3 |
| Bronchodilator | Albuterol (Inhaler), inhaler | 2 |
| Stimulants | Methylphenidate (Concerta, Ritalin),<br>Dexmethylphenidate (Focalin) | 7 |
| Sedative | Clonidine | 1 |
| Antihistamine and Antacid | Famotidine (Pepcid) | 1 |
| Laxative | Polyethylene Glycol 3350 (Miralax) | 1 |
| Supplements | Probiotics, Vitamins, Melatonin | 3 |

**Table S2.** Measures with Missing Observations.

| **Measure** | **Number of Missing Observations** |
| --- | --- |
| YSR-baseline | 2 |
| YSR-follow-up | 6 |
| ELS stress sensitivity | 3 |
| Parent Education | 6 |
| BMI baseline | 3 |
| Tanner follow-up | 5 |

YSR = Youth Self Report; ELS = Early Life Stress. Missing values were imputed using Multivariate Imputation by Chained Equations

**Table S3.**
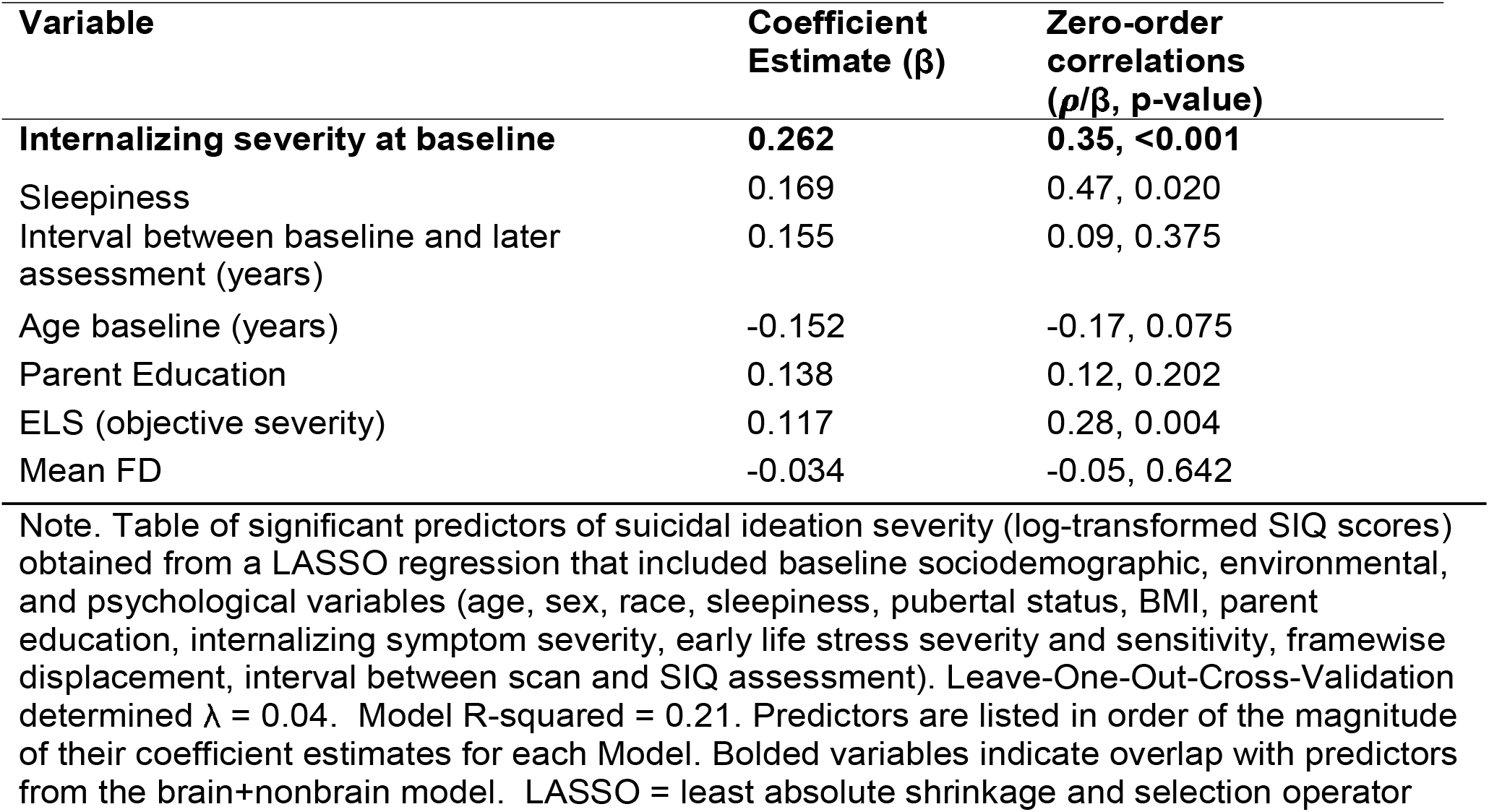
Nonbrain Predictors of Suicidal Ideation Severity from LASSO Regularized Regression (N=108)

| Variable | Coefficient Estimate ($\beta$) | Zero-order correlations ($\rho/\beta$, p-value) |
| --- | --- | --- |
| **Internalizing severity at baseline** | **0.262** | **0.35, <0.001** |
| Sleepiness | 0.169 | 0.47, 0.020 |
| Interval between baseline and later assessment (years) | 0.155 | 0.09, 0.375 |
| Age baseline (years) | -0.152 | -0.17, 0.075 |
| Parent Education | 0.138 | 0.12, 0.202 |
| ELS (objective severity) | 0.117 | 0.28, 0.004 |
| Mean FD | -0.034 | -0.05, 0.642 |

Note. Table of significant predictors of suicidal ideation severity (log-transformed SIQ scores) obtained from a LASSO regression that included baseline sociodemographic, environmental, and psychological variables (age, sex, race, sleepiness, pubertal status, BMI, parent education, internalizing symptom severity, early life stress severity and sensitivity, framework displacement, interval between scan and SIQ assessment). Leave-One-Out-Cross-Validation determined $\lambda = 0.04$ . Model R-squared = 0.21. Predictors are listed in order of the magnitude of their coefficient estimates for each Model. Bolded variables indicate overlap with predictors from the brain+nonbrain model. LASSO = least absolute shrinkage and selection operator

**Table S4.** Correlations between Baseline Participant Characteristics and Severity of SI.

| **Variable** | **Estimate ($\rho/\beta$)** | **p-value** |
| --- | --- | --- |
| Sex | 0.26 | 0.176 |
| Race (0=White, 1=POC) | -0.16 | 0.423 |
| Age baseline (years) | -0.17 | 0.075 |
| Tanner at baseline | -0.02 | 0.854 |
| **Internalizing severity at baseline** | **0.35** | **<0.001** |
| Interval between baseline and later assessment (years) | 0.09 | 0.375 |
| **ELS (objective severity)** | **0.28** | **0.004** |
| ELS (sensitivity) | 0.02 | 0.861 |
| Parent Education | 0.12 | 0.202 |
| BMI | 0.05 | 0.607 |
| Mean FD | -0.05 | 0.642 |
| **Sleepiness** | **0.47** | **0.020** |

Note. Spearman's $\rho$ correlations were computed for continuous variables and single variable linear regressions were performed for categorical variables. POC = people of color; ELS = early life stress; FD = Framewise Displacement; p-values < .05 are bolded.

**Table S5.**
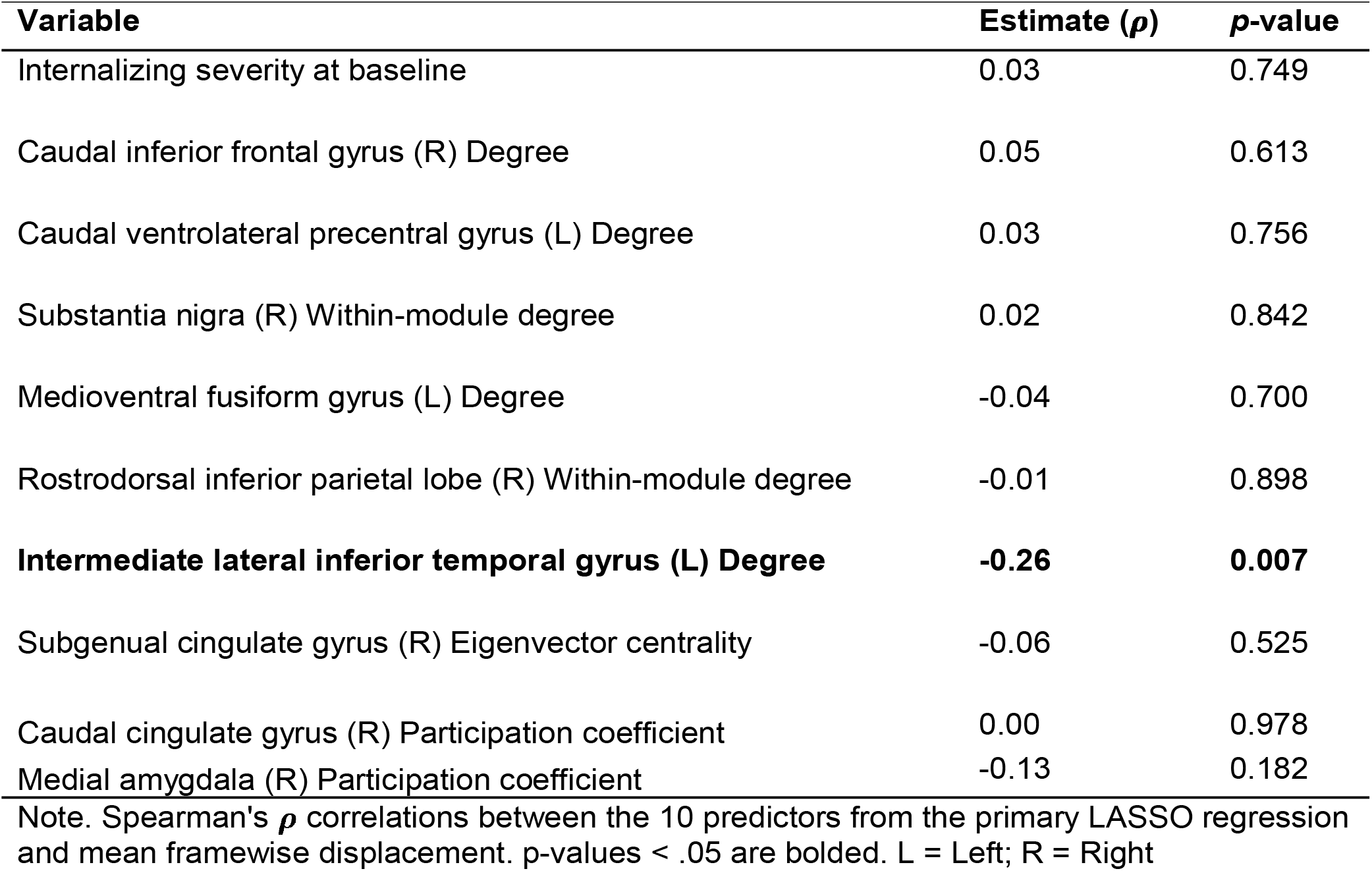
Correlations between Framewise displacement and LASSO-based predictors of severity of suicidal ideation.

This supplemental analysis assesses the correlation between average framewise displacement and the variables with non-zero estimates from the primary LASSO regression predicting severity of SI to examine whether levels of motion are correlated with the predictors of future SI severity. This analysis revealed only one significant correlation between degree of the left intermediate lateral inferior temporal gyrus and mean framewise displacement (P =-.26, *p=*.007). We therefore tested whether degree of the left intermediate lateral inferior temporal gyrus was associated with future severity of SI above and beyond mean framewise displacement. This analysis revealed that degree of the left intermediate lateral inferior temporal gyrus was associated with future severity of SI (β=0.25, *p=*.014) above and beyond mean framewise displacement (β=-0.03, *p=*.749).

**Figure S1.**
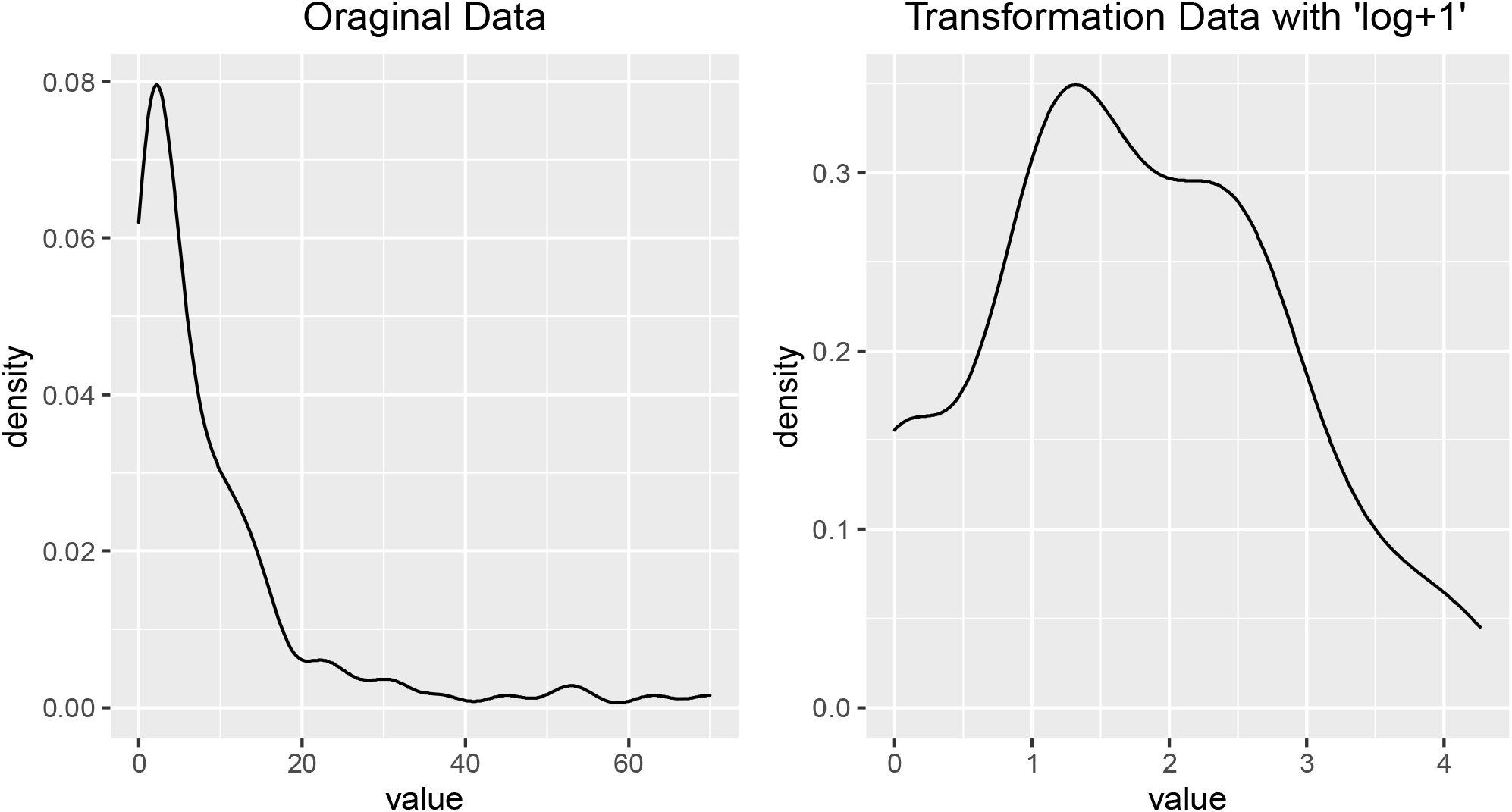
Log-transformation of suicidal ideation severity scores (N = 108). *Note:* 1 was added to scores given some values were zero.

